# ROLE OF COMMUNITY PROGRAMS IN SUSTAINING INTEGRATED HIV AND HEALTH SERVICES: A SCOPING REVIEW

**DOI:** 10.1101/2025.05.10.25327366

**Authors:** Ibrahim Bola Gobir, Piring’ar Mercy Niyang, Vera Labesa, Joy Musa, Bashir Zubayr, Itunu Dave-Agboola, Kolawole Olatunbosun, Terso Usha, Mabel Ikpeme, Pamela Gado, Onyeka Igboelina, Chika Obiora-Okafor, Dolapo Ogundehin, Deus Bazira

## Abstract

Community-centered initiatives are essential for tackling the persistent inequalities that perpetuate significant HIV-related disparities across various regions and countries worldwide. However, the lack of a systematic framework for analysis of the role of community-based programs in HIV prevention and ART monitoring impedes our ability to understand and study their effectiveness amidst scarce health professional resources. A scoping review was conducted to gather available evidence on the role of community programs in driving sustainable integrated HIV and health services, examining their impact on various interventions along the care continuum. Following JBI guidelines, a search was conducted for peer-reviewed literature published from January 2014 to December 2024. An initial search yielded 23 records. After screening for relevance and conducting cross-validation, 12 articles were selected. All the selected studies showed the positive impact of community programs on a wide range of aspects of the ART program. Thirty percent of articles (n = 3) highlighted the ability of community programs to mobilize resources and support, advocate for access to treatment, and monitor the delivery of antiretroviral therapy (ART) and related services, 20% (n=2) demonstrated that community programs can expand HIV/AIDS interventions and ensure a seamless continuum of services, from testing to care to treatment and the importance of community programs in empowering patients to take charge of their care, respectively. Findings from the scoping review underscore the integration of community programs in HIV and health services as a potentially effective strategy to address the growing shortage of health workers. Specifically, the review recommends, where possible, that decentralizing HIV services through community-based initiatives is a wise investment to increase access to essential health services, including ART

## Introduction

Community-centered initiatives are crucial for grasping and tackling the persistent inequalities that perpetuate significant HIV-related disparities across various regions and countries worldwide. (CDC, 2023). Community programs for HIV and health services can be interpreted and utilized in various ways, serving diverse purposes and holding different significance for various stakeholders, including program implementers, policymakers, and donors, each with distinct perspectives and interests. These programs encompass a wide range of initiatives and activities to address various aspects of the HIV epidemic within communities. This may include prevention programs, testing and counseling services, advocacy and awareness campaigns, support groups, peer education, and community mobilization. HIV community programs often involve collaboration between community organizations, healthcare providers, government agencies, and other stakeholders to address the diverse needs of communities affected by HIV/AIDS (Olubulyera, 2016; Japan International Cooperation Agency (JICA), 2021).

HIV community programs and community-based care (CBC) are related concepts but not necessarily the same. While both focus on addressing the needs of individuals and communities affected by HIV/AIDS, not all HIV community programs focus on delivering healthcare services directly. Some may focus on advocacy, education, or mobilization efforts, while others may include elements of community-based care within their programming. Conversely, community-based care programs may focus primarily on healthcare delivery and support services, with less emphasis on broader community engagement and mobilization. Hence, it is appropriate to assert that CBC offers substantial support throughout the care process, including all stages from infection to end-of-life care and attending to diverse facets of the care continuum. It also includes a variety of activities connected to AIDS that take place outside of conventional healthcare settings, although it may collaborate with official health and social agencies (Rachlis et al., 2013).

From the foregoing, the pivotal role of community programs in the global fight against HIV/AIDS cannot be overstated. These programs represent a beacon of hope, embodying grassroots efforts to combat the epidemic at its very core.

Decentralizing HIV services through community-based approaches has been used in many regions with limited healthcare resources (Rachlis et al., 2013; Bulstra, 2021), and with the scale-up of global HIV prevention and treatment came new challenges involving providing access to essential treatment for those who require it and offering ongoing long term care and support from individuals who are benefiting from life-prolonging therapies, recognizing the fact that HIV/AIDS is disease that comes with time (Rachlis et al., 2013). To maximize access to HIV prevention and treatment services in the presence of scarce professional health resources, health planners must sustain or perhaps improve on renewed interest in the potential role of community-based programs in HIV prevention and ART monitoring.

### Dynamics of global HIV statistics and control progress

In 2022, UNAIDS reported that approximately 39 million individuals worldwide were living with HIV. This total includes 37.5 million adults and 1.5 million children under 15 years of age, with women and girls comprising 53%. Regionally, Eastern and Southern Africa had the highest prevalence, with 20.8 million cases, followed by Western and Central Africa, which accounted for 4.8 million cases. (UNAIDS,2023; U.S Department of Health and Human Services, 2023). In 2022, approximately 1.3 million people worldwide contracted HIV, which marks a 38% reduction in new infections since 2010 and a 59% decrease since the 1995 peak. Women and girls made up 46% of all new HIV cases reported in 2022. (U.S. Department of Health & Human Services, 2023; UNAIDS, 2023).

In terms of access to HIV and health services, 84% of people with HIV worldwide have been tested and know their HIV status, and in 2022, approximately 86% of people living with HIV globally were aware of their status, leaving 14% (5.5 million individuals) unaware of and in need of testing services, (UNAID, 2023). This falls short of WHO’s 2025 target of 95% global HIV status awareness (WHO,2023). Knowing one’s HIV status through testing is a crucial entry point to a range of vital services, including prevention measures, treatment options, care programs, and support resources, all of which are essential for managing and living with the condition. As of the end of 2022, 29.8 million people with HIV (76% of all people with HIV) were accessing antiretroviral therapy (ART) globally, including 82% of pregnant women (UNAIDS, 2023). Individuals living with HIV who are aware of their status adhere to prescribed antiretroviral therapy (ART) and achieve an undetectable viral load can enjoy long and healthy lives.

Moreover, they will not transmit HIV to their HIV-negative sexual partners, a concept often referred to as "undetectable equals untransmittable" or U=U (U.S. Department of Health & Human Services, 2023). Ensuring awareness of HIV status is vital in sustaining the HIV care continuum, which represents the comprehensive journey of a person living with HIV, spanning from initial diagnosis to ongoing treatment and ultimately achieving an undetectable viral load. These steps involve receiving an HIV diagnosis, connecting with medical services, initiating ART, consistently following the treatment plan, and achieving viral suppression.

Enhanced access to HIV and health care services has shown a significant 69% decline in AIDS-related fatalities since the peak in 2004. Notably, in 2022, the number of people who died of AIDS-related illnesses worldwide was approximately 630,000, a substantial reduction from 2 million in 2004 and 1.3 million in 2010 (UNAIDS,2023). Despite these improvements, many individuals living with HIV or vulnerable to infections continue to face barriers in accessing essential services, including preventive measures, medical care, and treatment, and disappointingly, a cure for HIV remains elusive.

### Access to equitable quality HIV care

There has been inequality in the progress of reducing new HIV infections, increasing access to treatment, and ending AIDS-related deaths, with many vulnerable people and populations left behind (U.S. Department of Health & Human Services, 2023). Similarly, the COVID-19 pandemic has revealed how other competing wide-spread global epidemics and pandemics may negatively impact HIV treatment and prevention efforts, thereby disrupting essential services, including a surge in gender-related issues such as gender-based violence and teenage pregnancies and increased economic burdens. Furthermore, persistent HIV-related stigma, combined with existing social inequalities and exclusion, poses significant obstacles to progress, and if commitment and strong partnership are not maintained, the global response to HIV/AIDS may be at risk of being undermined. As noted by Boakye et al. (2024), stigma emerges as an amplifier of transmission risks and obstacles across the care continuum among key populations, with criminalization and social marginalization exacerbating barriers. Across the globe, key populations, including men who have sex with men and bisexual men, transgender women, female sex workers, and people who inject drugs, remain disproportionately burdened by HIV, facing infection rates dozens of times higher than the general population (Avert, 2024; Mayer & Allan-Blitz, 2019), and with the trend particularly evident in high- and middle-income countries (Korenromp et al., 2024). Thus, stigma and discrimination create immense, complex barriers, preventing further control of the disease, especially among marginalized populations. Of the 650,0000 global deaths from AIDS-related illnesses in 2021 (UNAIDS, 2022), marginalized populations accounted for the highest proportion (Halkitis et al., 2013; Spieldenner & Nieto, 2024).

Initiatives successfully countering stigma have included peer navigation services, mobile clinics, telehealth expansion, and advocacy campaigns. To successfully end HIV/AIDS by 2030 as targeted, response strategies must keep evolving to meet the marginalized where they are and to link all lives worth saving to the care required to save them. This would require decentralization of HIV services through community-based approaches, legal protections, and community empowerment efforts centered on affected populations’ leadership, especially in many regions with limited healthcare resources (Rachlis et al., 2013; U.S. Department of Health & Human Services, 2023; UNAIDS, 2023, Boakye et al., 2024).

### Rationale for the review

This review aims to identify community programs’ role in driving sustainable integrated HIV and health services, examining their impact on various interventions along the care continuum: diagnosis, linkage, treatment and retention, and other healthcare-related interventions such as mental health, sexual and reproductive health. Similarly, their impact on resource allocations, cost-effectiveness, efficiency, and value for money will be examined. Studies have consistently shown that integrating HIV services with other healthcare services has led to better outcomes than vertical programs (Bulstra, 2021). The overall body of evidence confirms the value and practicality of integration in enhancing the long-term sustainability of HIV programs and driving progress in the fight against HIV. This suggests that implementing more comprehensive and integrated ART programs could significantly improve the overall experience and healthcare outcomes of people living with HIV, leading to more effective and patient-centered care. Ensuring the long-term sustainability of HIV/AIDS healthcare services requires their integration into community programs.

### Methodology

Comprehensive literature search was carried out to determine the roles of community programs in driving sustainable integrated HIV and health services using a five-step methodology of the Joanna Briggs Institute (JBI).

### Literature Search Strategy

The search was set out to provide a comprehensive review of scientific literature highlighting the contribution of community programs to sustainable integrated HIV and health services. To determine the roles of community programs in driving sustainable integrated HIV and health services, a scoping review of published works in the last decade is conducted using the Joanna Briggs Institute (JBI) guidelines (Pollock et al., 2021). As scoping reviews are an invaluable form of evidence synthesis that addresses a particular research question, the overall goals of this review, therefore, are to gather available evidence on the roles of community programs in driving sustainable integrated HIV and Health Services and to identify key concepts and definitions, providing research frameworks or providing background, or contextual information on the phenomena of interest that provide actionable insight that can directly inform and improve policy decision, health care practices, and research priorities.

The review adhered to a five-step methodology of the JBI, which entailed formulating a research question, systematically identifying and searching for relevant studies, applying predetermined criteria to select studies for inclusion, extracting and organizing data from selected studies (charting the data), synthesizing and summarizing the findings (Zachary et al., 2023). The review was guided by the question: “What roles can community programs play in sustaining integrated HIV and health services?” The literature search was conducted using PubMed/MEDLINE, Google Scholar, Science Direct, and Academia. Search terms included "HIV and health services community programs" OR "Community programs role in HIV and health services" OR "community-led AIDS" OR “community HIV service integration”, “HIV programs sustainability”, necessary to identifying research studies and policy reports on significant progress of HIV/ART-related community programs. Relevant and additional literature was identified by searching scholarly databases for publications, and institutional reports released by non-governmental organizations (NGOs) and academic research centers were also reviewed. The reference lists of reviewed publications were checked for other relevant reports and papers.

### Study selection and eligibility

Due to the paucity of scientific studies specifically examining the roles of community programs in sustaining integrated HIV and health services, this review adopts a broad approach by incorporating various research designs and community support categories, enhancing the comprehensiveness and relevance of the review’s findings. The review includes qualitative and quantitative research papers on the topic, recognizing that diverse methodological approaches offer distinct yet complementary insight on the same topic to harness the strength of each, providing a more comprehensive understanding. Only papers that were published in the English language were included in this study. The review consists of various types of studies, randomized controlled trials, comparison groups (including pre-test, and post-test design), retrospective cohort studies, descriptive studies, and qualitative studies for exploring experience, as well as all studies that measured or examined any aspect related to integrated HIV/ART services with community programs (including access, coverage, adherence, advocacy, viral suppression, and survival as well as healthcare financing, sustainability, and resource allocations) were included. Studies published from January 2014 – December 2024 from across the globe were included in this review.

This scoping review considered a broad perspective when assessing the roles of community programs in scaling up sustainable integrated HIV and Health Services. For inclusion, the studies had to comprise (1) a definitive HIV/AIDS program group as the study population, (2) a defined role/intervention in the ART program, and (3) fulfill these role(s) in an organized manner that included a wide range of community programs functions (e.g., prevention programs, testing, and counseling services, advocacy and awareness campaigns, support groups, peer education, and community mobilization efforts, etc). These broad criteria were chosen to capture all available evidence on the roles of community programs in sustaining integrated HIV and health services. Thus, community-based ART programs that lacked a clear community-based support component were excluded from this study.

### Study selection

Based on the inclusion and exclusion criteria, 12 full-length papers were included because they provided valuable insight and relevant information on the roles of community programs on ART outcomes (appropriateness of study design, data collection methods, sampling strategy, and analytic approach as well as a sufficient description of the context according with th methodological requirements of a scoping review as specified by Peters et al. (2020). This resulted in 12 full-text articles assessing the roles of community programs in sustaining integrated HIV and health services, as illustrated in Figure 1

**Fig1:**
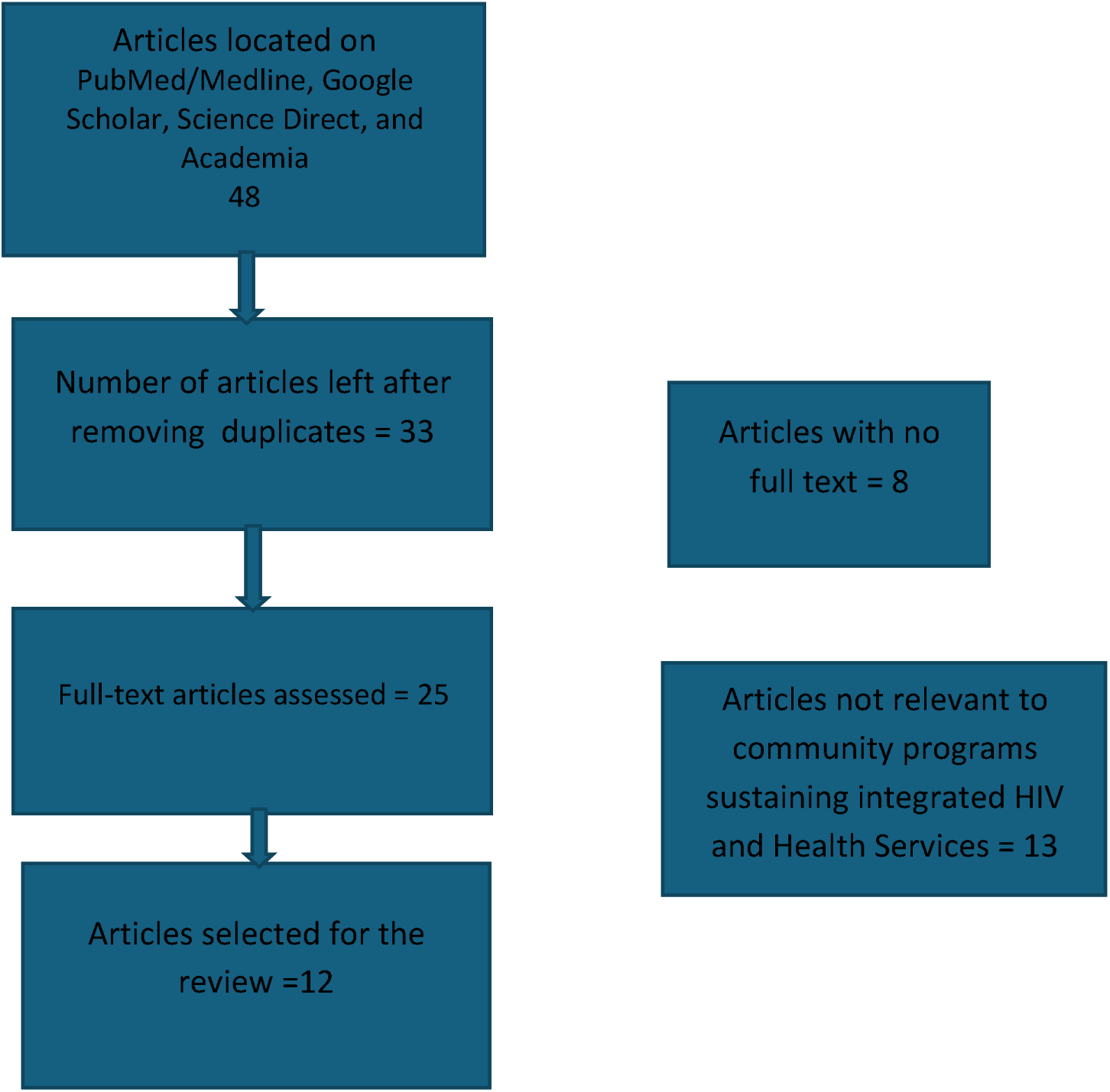
Flowchart of the study selection process Analysis. An analysis of outcomes of different functions of community programs around the globe, as reported in the 12 selected studies, was conducted to produce a concise summary of the roles of community programs in sustaining integrated HIV and health, as shown in Table 1.

## Result

### Thematic analysis of research findings Study design and population

The included articles in the final analysis reported on the outcomes of different functions of community programs around the globe. Most papers were published in the last decade (10 Years).

To have a robust assessment of the impact of community programs on sustainable integrated HIV and health services, the included studies used a wide range of methodological designs - descriptive studies, randomized controlled trials, quasi-experiments, retrospective/observational cohort studies, and qualitative studies. Additionally, the chosen studies encompass a diverse array of community programs that are involved in ART-related activities, and the study population ranged from the general population to specific key and priority populations such as sex workers, gays/bisexuals, and youths.

### The supportive role of the community

All the selected studies showed the positive impact of community programs on a wide range of aspects of the ART program. Firstly, the studies reviewed indicated that community programs could aid in expanding access to and increasing the coverage of ART programs through mobilization, advocacy, and monitoring (Reza-Paul et al., 2008; Argento, 2016; Baptiste et al., 2020). Moreover, the services provided by these community programs have been shown to increase adherence levels (Nachega et al., 2016). Furthermore, seven studies indicated that the community programs significantly improved the ART program’s testing, care, and treatment outcomes through community empowerment (Nglazi et al., 2011; Smith et al., 2015; Kerrigan, 2015). Finally, the summarized results in Table 1 also reveal that community programs can improve ART monitoring (Wouters et al, 2012).

**Table 1:** Data Extraction Table.

| Year | Authors | Method | Population | Intervention/role | Outcomes and Key Findings |
| --- | --- | --- | --- | --- | --- |
| 2014 | Zeng, W. et al | Data envelopment analysis | Rural health centers in Rwanda | Community-based health insurance (CBHI) and performance-based financing | Results show that health centers performance increased by 15.6% from 2006 through 2007. When the services are examined separately, each 1% growth of CBHI use was associated with 3.7% more prevention of mother-to-child transmission and 2.5% more voluntary counseling and testing services. |
| 2015 | Hoske, S.G. et al. | Mixed study through quantitative and qualitative study design | Opinion Leaders (OLs), Gay – youths | Community empowerment | Overall exposure to the intervention increased across waves. Statistically significant ($p < .05$) declines were observed for multiple sexual partners, condomless anal intercourse with any male partners, and with male partners of unknown HIV status. HIV stigma declined as well, but the trend was not statistically significant. |
| 2015 | Garcia, J. et al. | Community-based ethnography | Black men who have sex with men | Prevention | The study's results showed that men's relationships with social and public institutions |
|  |  |  | (BMSM) |  | are marked by fear and mistrust, making them more vulnerable to HIV. This mistrust discourages them from accessing and using essential healthcare services. However, the study's ethnographic findings highlight the potential of "safe space" as a crucial, yet underutilized component of HIV prevention efforts. |
| 2015 | Reza-Paul S, et al. | Prospective/<br>longitudinal | Sex workers | Mobilization/advocacy/monitoring | Program data show complete coverage of community outreach and greater than 90% clinic attendance for quarterly checkups by 2010. Reported condom use with the last occasional client increased from 65% to 90%. Surveys documented a halving of HIV and syphilis prevalence between 2004 and 2009, while gonorrhea declined by 80%. Between 2005 and 2013, clinic checkups tripled, whereas the number of STIs requiring treatment declined by 99%. New HIV infections also declined, and the primary study setting achieved strong cascade outcomes for HIV testing, antiretroviral treatment linkage, and retention. |
| 2016 | Smith, D.K. et al | A web-based descriptive survey | HIV Community – Based Organizations | Evaluating the CBO engagement with biomedical prevention services for HIV/AIDS | Results indicated Clinical CBOs reported higher budgets on average than nonclinical CBOs. The average of the reported proportion of clients served, by transmission risk group, were 41% MSM (men who have sex with men), 26% heterosexual females, 23% heterosexual males, 14% persons who inject drugs, 4% WSW (women who have sex with women), and 5% transgender persons. Nearly all CBOs reported providing HIV testing (92%), individual (86%), and group behavioral (80%) risk-reduction interventions, and linkage to social services (96%). However, more than half of clinical CBOs, compare to <5% of nonclinical CBOs reported providing clinical services. Clinical CBOs were also significantly more likely to provide treatment for opiate addiction (33% vs. 0%), linkage to social services (100% vs. 90%), and partner services (90% vs. 77%). |
| 2016 | Nachega et al. | Observational and experimental studies with primary data using cross-sectional, case-control, and cohort (prospective and retrospective) and randomized controlled trial (RCT) designs. | HIV-infected individuals initiated on ART. | Community-based ART delivery | Results showed comparable outcomes in terms of ART adherence (RR = 1.02, 95 % CI 0.99 to 1.04), virologic suppression (RR = 1.00, 95 % CI 0.98 to 1.03), and all-cause mortality (RR = 0.93, 95 % CI 0.73 to 1.18). The result of a pooled analysis from the RCTs (RR = 1.03, 95 % CI 1.01 to 1.06) and cohort studies (RR = 1.09, 95 % CI 1.03 to 1.15) found that participants who received community-based interventions showed significantly higher rates of treatment engagement compared to those who did not. |
| 2011 | Nglazi M.D., et al. | Prospective/longitudinal | People living with HIV | Testing/care/Treatment (1<sup>o</sup>-3<sup>o</sup> roles) | The treatment was highly effective, with 93% of patients achieving viral suppression after 16 weeks. Moreover, the consistent viral suppression rate observed over consecutive years of recruitment indicates that patients maintained good adherence to their treatment regimens, resulting in sustained viral control. |
| 2012 | Wouters et al. | Systematic review | PLWHA, lay members of the community | ART monitoring | Findings reported an unambiguously positive impact of community support on a wide range of aspects, including access, coverage, |
|  |  |  | (community health workers, community care workers, lay health workers, treatment buddies, field officers, peer educators/counselors , adherence supporters.) |  | adherence, virological and immunological outcomes, patient retention, and survival. The review indicates that community support initiatives address five often cited challenges to ART scale-up: the lack of integration of ART services into the general health system; the growing need for comprehensive care; patient empowerment; defaulter tracing; and the crippling shortage in human resources for health. Therefore, a well-planned and well-executed community support initiative is essential for a successful and sustainable public sector ART program. |
| 2015 | Smith et al. | Individual-based mathematical model | Asymptomatic HIV patients | Home HTC | The model predicted the implementation of home HTC in addition to current practice to decrease HIV-associated morbidity by 10–22% and HIV infections by 9–48% with increasing CD4 cell count thresholds for antiretroviral therapy initiation. Community-based HTC with improved linkage can result in widespread HIV testing and treatment, ultimately reducing the number of people living with HIV-related illnesses and deaths. |
| 2015 | Kerrigan D. et al | Lit review | Sex workers | Community Empowerment | When community empowerment increases, there's a significant decrease in the risk of contracting HIV, gonorrhea, chlamydia, and syphilis. Specifically, the odds of getting infected with these sexually transmitted infections (STIs) drop by 32% for HIV, 39% for gonorrhea, 26% for chlamydia, and 47% for syphilis. Additionally, community empowerment is linked to a 227% increase in consistent condom use with clients, indicating a positive impact on safe sex practices. |
| 2016 | Argento E, et al. | Prospective/ longitudinal | Sex workers | Community mobilization | When sex workers feel a stronger sense of community and connection with each other (perceived social cohesion), they are more likely to experience a decrease in clients refusing to use condoms, regardless of other factors. |
| 2020 | Baptiste S. et al. | Lit review | People living with HIV | Mobilization/ advocacy/monitoring | Community-led monitoring resulted in notable enhancements in healthcare, such as enhanced<br>healthcare service accessibility, improved health outcomes, reduced waiting times, strengthened community partnerships, decreased occurrences of important medicine and supply shortages, and increased HIV testing and viral load testing.<br>Specifically, within 18 months: ART stockouts decreased by 8.4%, Viral load testing stockouts decreased by 10.7%, 23,618 more people started antiretroviral therapy (ART), 16,844 more viral load tests were performed, Viral suppression rates increased by 29% and average quality of care ratings improved from 3.8 to 4.2 out of 5 across all health sites. |

### Community mobilization, advocacy, and monitoring

Out of 10 studies examining community programs’ role in integrated HIV and health services, 3 studies (30%) highlighted the ability of community programs to mobilize resources and support, advocate for access to treatment, and monitor the delivery of antiretroviral therapy (ART) and related services. (Reza-Paul et al., 2015; Argento, 2016; Baptiste et al., 2020). A study by Baptiste et al. (2020) showed that community-led monitoring could result in increased access and utilization of services, improved health, decreased mortality, reduced waiting times, enhanced community relationships, earlier initiation of antiretroviral treatment, infrastructure upgrades, reduced stock-outs, increased HIV testing, and increased use of viral load testing in treatment monitoring. In this way, community support initiatives can – as shown by Nachega et al. (2016) and Garcia et al. (2015) – increase treatment adherence and engagement with related services that consequently lead to viral suppression by connecting patients to essential healthcare services and programs (TB, HIV/AIDS), promoting safe spaces as social support, that address stigma without compromising the quality of care.

### Scaling up of testing, Care and Treatment

Among the articles discussing the role of community programs in scaling up antiretroviral therapy (ART), 2 articles (20%) demonstrated that community programs can expand HIV/AIDS interventions and ensure a seamless continuum of services, from testing to care to treatment. (Nglazi et al., 2011; Smith et al., 2015). In this way, community support initiatives meet the emerging and overwhelming needs associated with chronic AIDS care by making community-based HIV testing and counseling more accessible and effective, allowing individuals to receive diagnosis and support earlier in their infection journey, even before they typically seek facility-based testing and care.

### Empowered ART patients

The complex nature of HIV/AIDS requires a patient-centered approach that considers individuals’ capacity to manage their condition. Two studies (Hoske et al., 2015; Kerrigan, 2015) highlighted the importance of community programs in empowering patients to take charge of their care. Specifically, Kerrigan (2015) found that sex workers who participated in community programs developed essential self-management skills, enabling them to Make informed decisions about their health and treatment, advocate for their needs and rights, and negotiate with healthcare providers for quality care. This approach emphasizes the critical role of community programs in enhancing patients’ autonomy and ability to navigate the healthcare system effectively. Through such community empowerment, they experienced a decline in HIV, gonorrhea, chlamydia, high-titer syphilis, and other sexually transmitted infections through increased consistent condom use with their clients.

### Access and equitable healthcare

As is the case, most funding for HIV/AIDS services is used to subsidize providers directly. Thus, most HIV/AIDS services, such as ART, PMTCT, and VCT, are provided free of charge to the patient in many recipient countries. However, these services, which are highly influenced by the recommendations of providers, were improved by Community-Based Health Insurance (CBHI), suggesting an alternative way to finance HIV/AIDS services efficiently and sustainably (Zeng et al., 2014). Improvement in the efficiency of HIV/AIDS services by CBHI underscores the fact that investing resources in health insurance would yield multifaceted benefits, not only relieving the resource constraints on HIV/AIDS with improved efficiency but also facilitating the services delivery with increased demand. Allowing CBOs to inform us about their interest and resource needs to take PrEP, PEP, and TasP from research into practice so that they can effectively integrate support for clients using these biomedical interventions in their HIV prevention programs is another efficient means of engendering access and equitable healthcare for HIV. CBOs with clinical capacity are positioned to provide PrEP, nPEP, and TasP care to their clients directly but are not currently meeting the perceived need because of limited financial, training, and client information resources. Even the CBOs that do not currently offer clinical services, as Smith et al. (2016) noted, are nevertheless interested in providing education, referral, and linkage to clinical care elsewhere, as well as other support services. HIV/AIDS financing in this regard should now be advocated. Additionally, underserved and marginalized PLHIV, such as KPs, will have the opportunity to receive quality care using community models that are explicitly focused on these populations. This will improve the uptake of HIV services among these populations, thereby reducing transmission. (Argento E, et al. 2016)

## Discussion

The role of community programs for sustainable and integrated HIV and health services differed most on the range of services provided as we noted heterogeneity regarding types of services provided over different periods along the continuum of care. Typical services offered include measures to prevent HIV transmissions, such as initiatives that promote risk reduction, strategies, education, and support to help individuals adopt safer behaviors and practices; HIV Testing and Counseling (HTC); Prevention of Mother-to-Child Transmission (PMTCT); ART provision and follow-up, including adherence monitoring; comprehensive medical and nursing care, including treatment of STIs and management of symptoms, prevention and treatment of opportunistic infections; nutrition support and food supplementation; palliative care; financial support in the form of income-generating activities (IGA) or community funds; material support, including technical support; information, communication, and education activities; and psychosocial support. Such heterogeneity underscores the multi-dimensional roles of community programs in HIV and health services.

Recent research published within the last decade highlights the growing importance of community programs in health systems research. All selected studies consistently showed that community programs positively impact various aspects of antiretroviral therapy (ART) programs. Specifically, community support was found to increase access to ART programs in resource-limited settings and expand coverage of ART programs. Overall, community programs play a crucial role in improving the effectiveness and reach of ART programs, particularly in areas with limited resources (Brien et al., 2010; Kerrigan et al., 2015). In addition, the services provided by these community programs have been shown to increase adherence levels (Wouters et al., 2012; Cowan et al., 2019; Baptiste et al., 2020).

Additionally, several studies showed that community-based support significantly enhances the effectiveness of ART programs, leading to better virologic outcomes, higher rates of viral suppression, and improved immunological outcomes, with greater immune system restoration. In other words, patients who received community support had better health outcomes, with higher viral suppression and immune recovery rates, than those without such support (Ngalazi et al., 2010; Xiao et al., 2012; Nnko et al., 2019). Finally, the results displayed in the literature grid also indicate that community programs can improve levels of patient retention (Wouters et al., 2012; Smith et al., 2015; Kerrigan et al., 2015; Chang et al., 2017; Brown et al., 2018) and increase rates of survival in ART patients in high HIV prevalence, resource-limited settings (Daudt et al., 2013; Cowan et al., 2019). More so, a comprehensive approach to ART can also focus on an individual’s strengths, resources, and ability to address the challenges of HIV/AIDS and ART. Two studies (Kerrigan et al., 2015; Cowan et al., 2019) applied a patient-centered approach to chronic disease care to investigate how community programs empower patients to manage their condition. Additionally, they identified loss to follow-up as the main reason for patient attrition and recognized that addressing this requires preventive and reactive strategies, including psychosocial support and regular home visits. Implementing these measures can minimize patient attrition, and patients can receive continuous care and support.

## Conclusion

The scoping review that included 12 studies has indicated that community programs integration in HIV and health services is a potentially effective strategy to address the growing shortage of health workers and to broaden care to accommodate the needs associated with chronic HIV/AIDS. Current research suggests that decentralizing HIV services through community-based initiatives is a wise investment to increase access to essential health services, including antiretroviral therapy (ART). Therefore, healthcare policymakers, managers, and providers should recognize the value of community programs in combating HIV/AIDS and work to enhance and support their role in the response to the epidemic.

## Data Availability

All data produced in the present work are contained in the manuscript

